# Access to Care in PANS: A Survey of Families’ Journeys to and Experiences with IVIG Treatment

**DOI:** 10.64898/2025.12.23.25342929

**Authors:** Denise Calaprice, Christina Moon, Melanie Helvick, Chase Harvey, Angela Tang, Kelsey Aguirre, James Hunt, Ryan Tererri, Christopher Whitty, Megan Fitzgerald

## Abstract

**Background:** Pediatric Acute-Onset Neuropsychiatric Syndrome (PANS) is characterized by abrupt-onset neuropsychiatric symptoms, often infection- or immune-triggered. Intravenous immunoglobulin (IVIG) is recommended by expert guidelines for select cases, yet insurance denials and high out-of-pocket costs have been major barriers to access.

**Methods:** We surveyed 60 caregivers and/or adult PANS patients who pursued IVIG therapy, collecting data on insurance experiences, treatment protocols, financial strategies, and patient/caregiver quality of life (QoL) before and after treatment.

**Results:** Most patients (88%) eventually received IVIG; 10% were still in pursuit, and 2% discontinued trying. Insurance approval without appeal occurred in only 18%, while many families faced multiple denials and appeals. Insurance approvals were more expeditious for patients with comorbidities for which IVIG is typically covered; among those receiving IVIG for PANS alone, only 5% were treated within a month of the doctor’s order (compared to 38% of those receiving IVIG for comorbidities as well as PANS) and 14% waited ≥9 months before treatment began. Financial strain was routine: one-third of families without substantial coverage (i.e., insurance that covered >=70% of expenses) reported extreme stress (10/10), 58% borrowed money, and 21% sold major assets. Even families with substantial insurance coverage commonly depleted savings or took on additional work. Prior to IVIG, patient Quality of Life (QoL) ratings were poor (means 2.1–2.8 across domains), with over one-third selecting the lowest possible ratings. During the six months following IVIG initiation, mean scores rose to 6.2–6.8, with over 60% reporting “good” to “exceedingly good” outcomes. Caregivers reported parallel gains, with family QoL ratings rising from 2.4–4.0 pre-treatment to 5.7–6.6 post-treatment.

**Conclusions:** Families pursuing IVIG for PANS faced prolonged delays, repeated denials, and extreme financial strain, often resorting to loans, asset sales, and additional work. Despite these burdens, IVIG was associated with marked improvements in quality of life for both patients and caregivers, underscoring the treatment’s potential benefits and the urgent need for more equitable access.

## Introduction

Pediatric Acute-onset Neuropsychiatric Syndrome (PANS) is a rare and frequently disabling disorder characterized by the sudden onset of obsessive-compulsive symptoms and/or severely restricted food intake, together with additional cognitive, behavioral, and somatic symptoms that may include anxiety, irritability, depression, sleep disturbance, urinary frequency, and significant academic and functional regression^1^. Although the diagnostic criteria for PANS require that onset be acute, they do not require a specific type of triggering event—episodes may be triggered by a variety of infections, inflammatory events, or environmental exposures, or triggers may be unknown. A closely related, older diagnosis, Pediatric Autoimmune Neuropsychiatric Disorders Associated with Streptococcal infections (PANDAS) has a nearly identical clinical presentation but requires that the acute onset closely follow a Streptococcal infection^2^. Since children with PANDAS typically also have PANS, we use the term PANS throughout to encompass both PANS and PANDAS.

Mounting evidence supports the role of aberrant immune responses in PANS pathophysiology^3,4^. A variety of inflammatory markers are present at elevated rates^5^; and rates of immunological comorbidities, including both autoimmune conditions and immune deficiencies, are much higher than expected in both patients and first-degree relatives^6–8^. Imaging studies demonstrating localized abnormalities support a specific role for basal ganglia autoantibodies in the disease mechanism^9–11^. Given the compelling data supporting a basis in immune dysregulation, expert consensus guidelines recommend immunomodulatory treatment for PANS, often alongside psychiatric and behavioral interventions^12^.

Intravenous immunoglobulin (IVIG), a human-derived immune globulin product delivered by infusion, is the principal (and FDA- and EMA-approved) treatment for immunoglobulin deficiencies as well as a common treatment for neuroinflammatory conditions such as autoimmune encephalitis, Kawasaki’s disease, and polyneuropathy. Although many PANS patients respond to more conservative therapies, IVIG has emerged as a frequently-prescribed, albeit off-label, therapeutic option for those who do not. Current expert consensus guidelines recommend IVIG for moderate-to-severe cases, particularly those with evidence of immune dysfunction or failure to respond to first-line treatments^12^. IVIG is presumably helpful in such patients because of its compound actions in combatting the infection(s) that may trigger flares, reducing neuroinflammation, and suppressing autoantibody production. Despite the published expert recommendations, the coherent scientific rationale, and its frequent use, however, rigorous data regarding the efficacy of IVIG in PANS remain limited. Although some clinical trials have been conducted, results have been mixed and their interpretation limited by factors including lack of an appropriate outcome measure for this heterogeneous condition, restrictive eligibility criteria, and sample sizes too small to permit a full understanding of response heterogeneity^13^—all issues that are common in rare disease clinical research.

Unlike the more commonly-used, oral immunomodulatory agents, IVIG is extremely expensive, with typical costs ranging from $5,000 USD to over $20,000 USD per infusion depending on dosage and patient weight. The financial burden is often amplified by the need for repeated infusions to achieve satisfactory effect^14^. Insurance companies frequently deny coverage for prescribed IVIG in PANS, citing its off-label status and the paucity of clinical trials. Families whose children have failed more conservative approaches must then decide between forgoing this treatment recommended by their physician, appealing coverage denials, or seeking alternative sources of funding. Success in appeals and/or fundraising is not assured, and even when it is ultimately achieved, these measures often take considerable time—months or even years—leading to delays in treatment that can have profound consequences. Prolonged disease activity has been associated with more entrenched psychiatric symptoms, greater academic decline, and worsening family functioning^8,15,16^.

In response to these challenges, legislative initiatives in several U.S. states have sought to mandate insurance coverage for IVIG when prescribed for PANS, in keeping with expert guidelines. To date, however, no published data have addressed how families navigate obtaining IVIG for PANS, what the process typically involves, and whether the real-world outcomes are perceived to justify the costs and sacrifices. In this study, we surveyed families whose PANS children had been prescribed IVIG, some of whom succeeded in receiving insurance approval and some of whom did not. Our aim was to report on influences that contributed to the decision to pursue IVIG treatment, the types of specialists from whom orders were obtained, the insurance review processes and outcomes, the ultimate financial burdens and means of satisfying them, and the perceived functional and quality of life outcomes for both PANS patients and their families.

## Methods

### Study Design

This study utilized an online, cross-sectional survey to assess the economic, functional, and quality of life impacts of IVIG treatment among individuals who reported having been diagnosed by a healthcare practitioner with confirmed or strongly suspected PANS. Using two nearly identical survey iterations, administered on TrialKit and Qualtrics platforms respectively (due to a technology transfer), quantitative and qualitative data were obtained regarding clinical history, influences on the decision to pursue IVIG, types of specialists from whom orders were obtained, insurance review processes and outcomes, treatment received, direct and indirect financial burdens and means of satisfying them, and perceived function and quality of life before and after IVIG treatment for both the PANS patients and their families. To assess how these varied with IVIG treatment, many of the questions asked about experiences during three periods of time: the 3-month period prior to when IVIG was ordered by a medical professional; the “waiting period” between when IVIG was ordered and when administration actually began (of variable length, generally driven by insurance approvals and other financial considerations); and the 6-month period beginning with first IVIG administration. Data were collected between April 2024 and June 2025.

### Recruitment and Participants

Participants were recruited via targeted email outreach to multiple PANS/PANDAS-focused online communities. Advertisements were specifically crafted to avoid bias with respect to anticipated outcomes or treatment perspectives, stating simply that the researchers were seeking to understand families’ experiences. Inclusion criteria required participants to be age 7 or older, to reside in the U.S., to have a formal diagnosis or strong clinical suspicion of PANS (per patient or parent/guardian report), and to either have received or attempted to access IVIG treatment prescribed by a licensed clinician. Participants under the age of 18 were required to be assisted by a parent or guardian in completion of the survey, and those over 18 were instructed to consult their parents/caregivers about any questions relating to the time they were under their care. Upon participant request, Brain Inflammation Collaborative Clinical Research Associates (CRAs; CM and MH) were available to assist participants in understanding survey questions. In no case did the CRAs suggest how questions should be answered.

The protocol, survey instruments, recruitment materials, and informed consent and assent language and process were approved by WCG IRB (for the TrialKit iteration) and Sterling IRB (for the Qualtrics iteration). Informed consent was obtained electronically via the survey technology for all adult participants, including both adult patients and adult caregivers. Assent was obtained electronically for minors. If eligibility criteria were not actively affirmed and consent and assent (as applicable) obtained, the participant was automatically exited from the technology system without having had the ability to view or answer survey questions.

### Data Cleaning

Data review and cleaning were performed to remove any duplicate responses (based on a combination of email address and demographic traits) and any responses that were either too incomplete to be used (no information provided beyond demographics/baseline characteristics), or that appeared unreliable based on illogical content. For the TrialKit iteration, no responses required removal as part of this process; for the Qualtrics iteration 40 responses were initially obtained and 30 remained after data cleaning. The TrialKit and Qualtrics datasets were merged using a shared unique participant identifier (UniqueID), and the merged dataset contained a total of 60 responses. Data processing was conducted using R statistical software (version 4.1.2), utilizing the following packages: *dplyr* for data manipulation, *janitor* for data cleaning, *stringr* for text processing, *readxl* and *openxlsx* for data import, *purrr* for functional programming, and tibble for data structuring, *tidygeocoder*, for turning IP Addresses into broad regions, *stringr* for text conversions, and *tidyverse*.

### Data Analysis

Descriptive statistics, including means, medians, ranges, and proportions, were assembled using Microsoft Excel (Microsoft Office 365). A complete-case approach was applied, with no imputation for missing data. N’s vary across the tables and figures depending on the number of people who answered each question.

Subgroups were defined to facilitate description of key variables across participant characteristics. Regional subgrouping was determined based on participants’ self-reported locations. Regions were divided as follows: **Northeast**: CT, ME, MA, NH, NJ, NY, PA, RI, VT; **Midwest**: IL, IN, IA, KS, MI, MN, MO, NE, ND, OH, SD, WI; **South**: AL, AR, DE, District of Columbia (DC), FL, GA, KY, LA, MD, MS, NC, OK, SC, TN, TX, VA, WV; **West**: AK, AZ, CA, CO, HI, ID, MT, NM, NV, OR, UT, WA, WY. For participants who did not provide location information, general geographic locations were inferred from participant IP addresses at the time of survey completion. IP data were used solely to assign participants to the broad regions and were not linked to any personally identifying information. Insurance coverage was categorized as “none to minimal” if participants either reported receiving no coverage or indicated coverage of less than 10% of expenses. “Substantial coverage” was defined as reporting greater than 70% of treatment costs covered by insurance. No respondent fell between these two categories.

## Results

### Characteristics of Participants and Treatment

The PANS patients represented in this study were 50% male and 42% female at birth (8% not specified) and identified as 53% male, 40% female, and 3% transgender (5% not specified) (Table 1). Mean current age was 14.4 years and age at first IVIG treatment was 11.2 years. The majority of those completing surveys were parents, though 28% were the patients themselves. At the time of the survey, respondents were distributed across the U.S., with the largest portion in the Midwest (28%), followed by the West (25%), South (25%) and Northeast (20%). The majority of patients (75–9%) were insured under Preferred Provider Organization plans (PPO). Only a small number of subjects changed insurance providers during the process of obtaining and receiving IVIG treatment.

**Table 1.** Characteristics of Patients and Respondents.

|  |  |
| --- | --- |
| PANS Diagnostic Certainty | N=53 |
| Confirmed by a licensed medical professional | 53 (100%) |
| Suspected | 0 (0%) |
| Patient Sex at Birth | N=60 |
| Male, N (%) | 30 (50%) |
| Female, N (%) | 25 (42%) |
| Not specified, N (%) | 5 (8%) |
| Patient Sex Gender Identity | N=60 |
| Male, N (%) | 32 (53%) |
| Female, N (%) | 24 (40%) |
| Transgender, N (%) | 2 (3%) |
| Not specified, N (%) | 3 (5%) |
| Patient Age | N=60 |
| Mean | 14.4 |
| Median | 14 |
| Range | 6–30 |
| Patient Age at First IVIG | N=48 |
| Mean | 11.2 |
| Median | 10 |
| Range | 3–27 |
| Respondent relationship to patient | N=60 |
| Mother, N (%) | 23 (38%) |
| Father, N (%) | 2 (3%) |
| Parent (undefined), N (%) | 18 (30%) |
| Self, N (%) | 17 (28%) |
| Geographic Region | N=60 |
| Northeast, N (%) | 12 (20%) |
| Midwest, N (%) | 17 (28%) |
| South, N (%) | 15 (25%) |
| West, N (%) | 15 (25%) |
| Not specified and could not be determined, N (%) | 1 (2%) |
| Type of Insurance, before IVIG Order | N=48 |
| Health Maintenance Organization (HMO), N (%) | 4 (8%) |
| Preferred Provider Organization (PPO), N (%) | 36 (75%) |
| State Health Plan (e.g. Medicaid), N (%) | 4 (8%) |
| Uninsured, N (%) | 2 (4%) |
| Type of Insurance, after IVIG Order | N=48 |
| Health Maintenance Organization (HMO), N (%) | 7 (15%) |
| Preferred Provider Organization (PPO), N (%) | 38 (79%) |
| State Health Plan (e.g. Medicaid), N (%) | 4 (8%) |
| Uninsured, N (%) | 1 (2%) |

Eighty-eight percent of the patients surveyed eventually did receive IVIG treatment; 10% were still in pursuit; and 1 family had discontinued the quest (Table 2). Dominant influences on the decision to pursue IVIG included personal research (mean rating=9.0/10; 86% of respondents endorsing ratings of 8, 9, or 10) and patient testimonials (mean=8.9; 85% rating 8, 9, or 10). Recommendations from professionals (mean 8.7) and outcomes from previous treatments (mean=8.0) also played key roles, while factors like insurance/financial considerations, support group inputs, and side effect concerns were less influential.

**Table 2.**
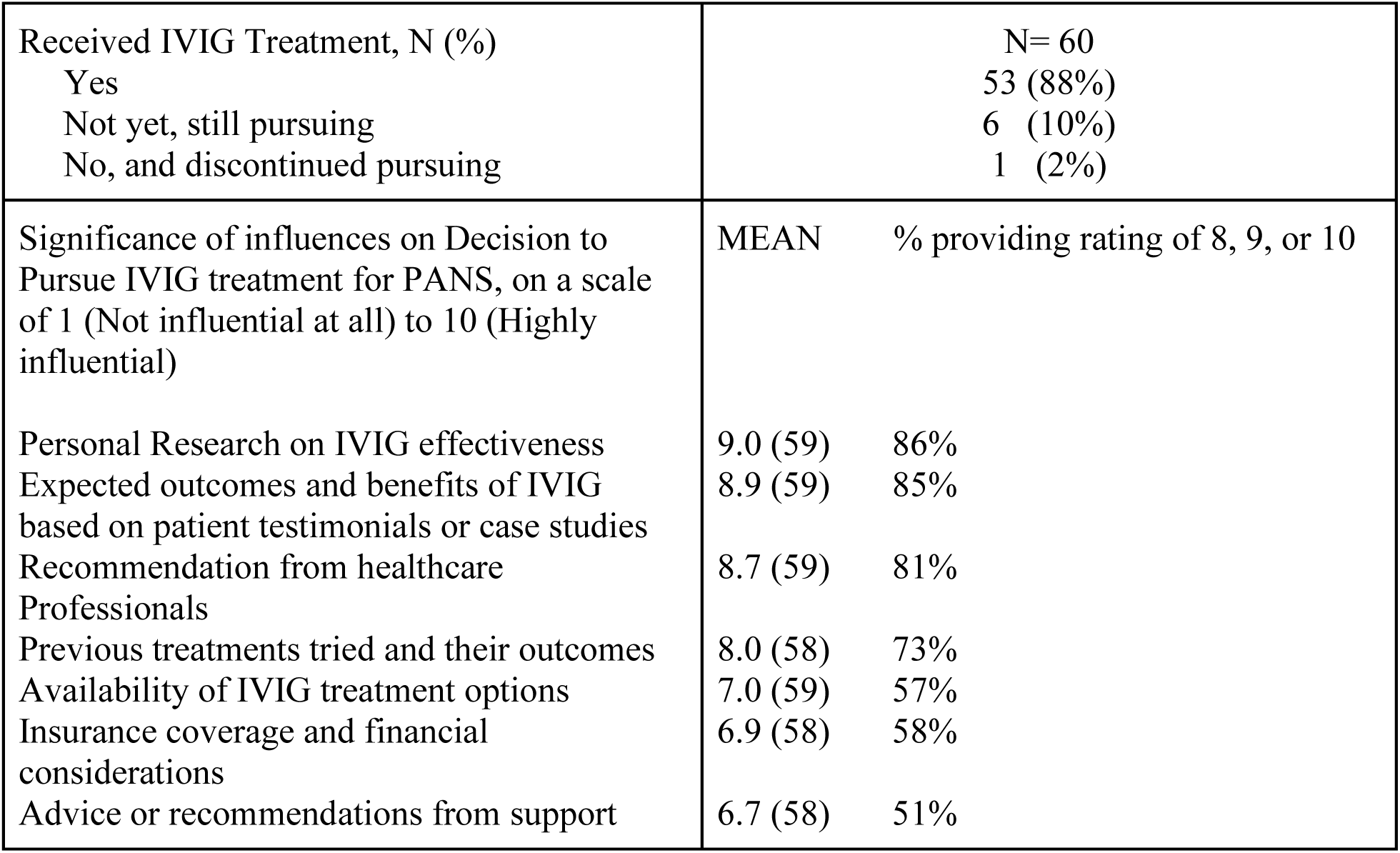
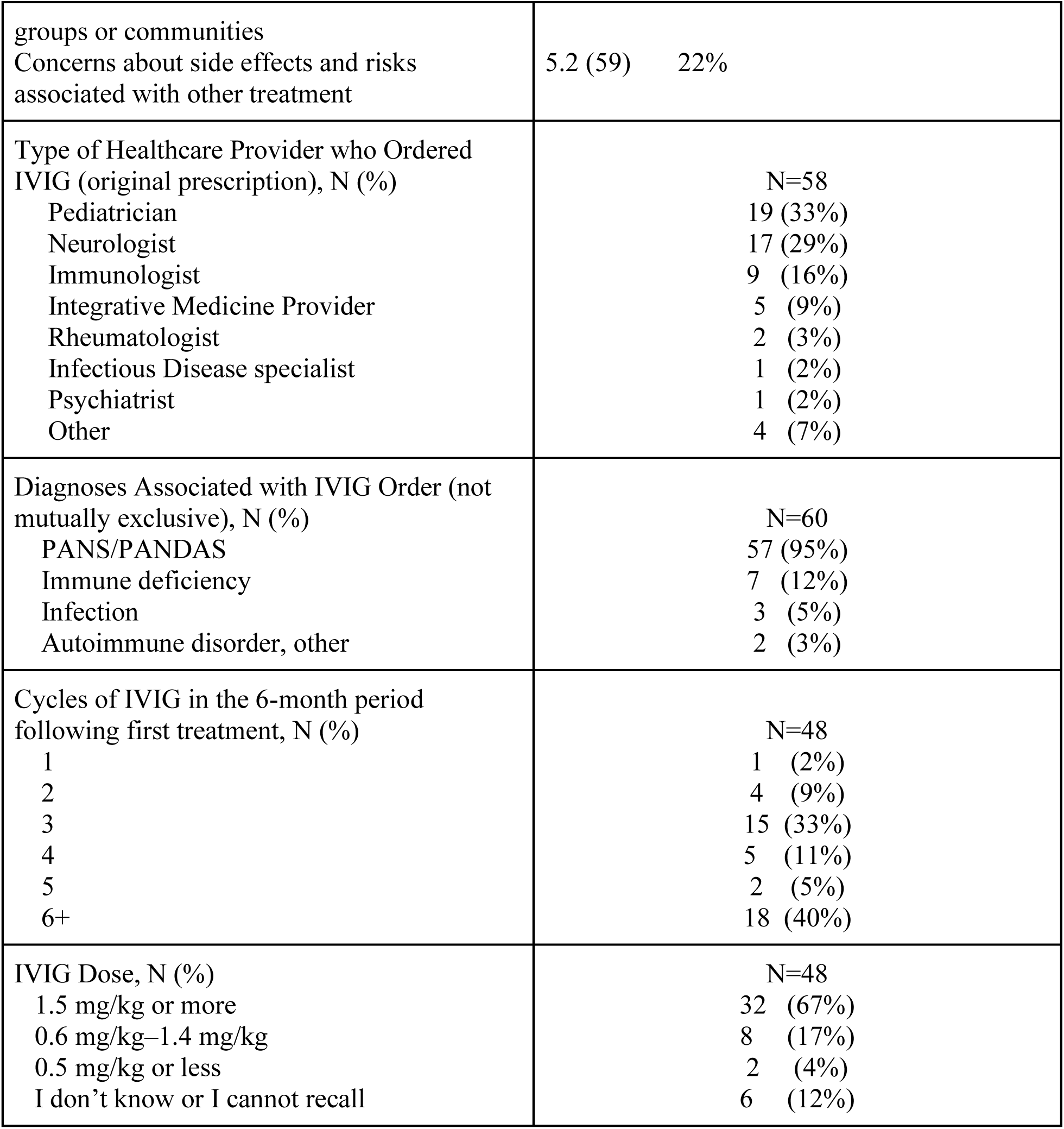
Characteristics of IVIG Treatment.

For most patients (95%), IVIG was ordered specifically for PANS. For a few, however, IVIG was also, or alternatively, ordered for immune deficiency (12%) or other autoimmune or infectious conditions. Pediatricians (33%), neurologists (29%), and immunologists (16%) were the most frequent prescribers. Treatment regimens varied; the most common (40%) involved receipt of six or more IVIG cycles in the first six months post-initiation. Most patients (67%) received doses ≥1.5 g/kg, and 89% received at least three treatment cycles.

### Insurance Coverage and Appeals

In most cases, insurance coverage for IVIG was initially denied (Figure 1; note that N=17 because this question was only asked explicitly in the Qualtrics iteration of the survey). Only 18% of the families surveyed received approval without appeals, of whom one-third submitted IVIG claims for a non-PANS diagnosis. More than three times this number required at least two appeals before coverage was obtained and an equal number were denied even after all possible appeals were exhausted. Actions described in the open-ended comments field to this question included filing for expedited review (N=1), reaching out to employers (N=2) or congressional representatives (1), threatening legal action (N=1), participating in a research study (N=1), or changing insurance (N=1), before coverage was obtained.

**Figure 1.**
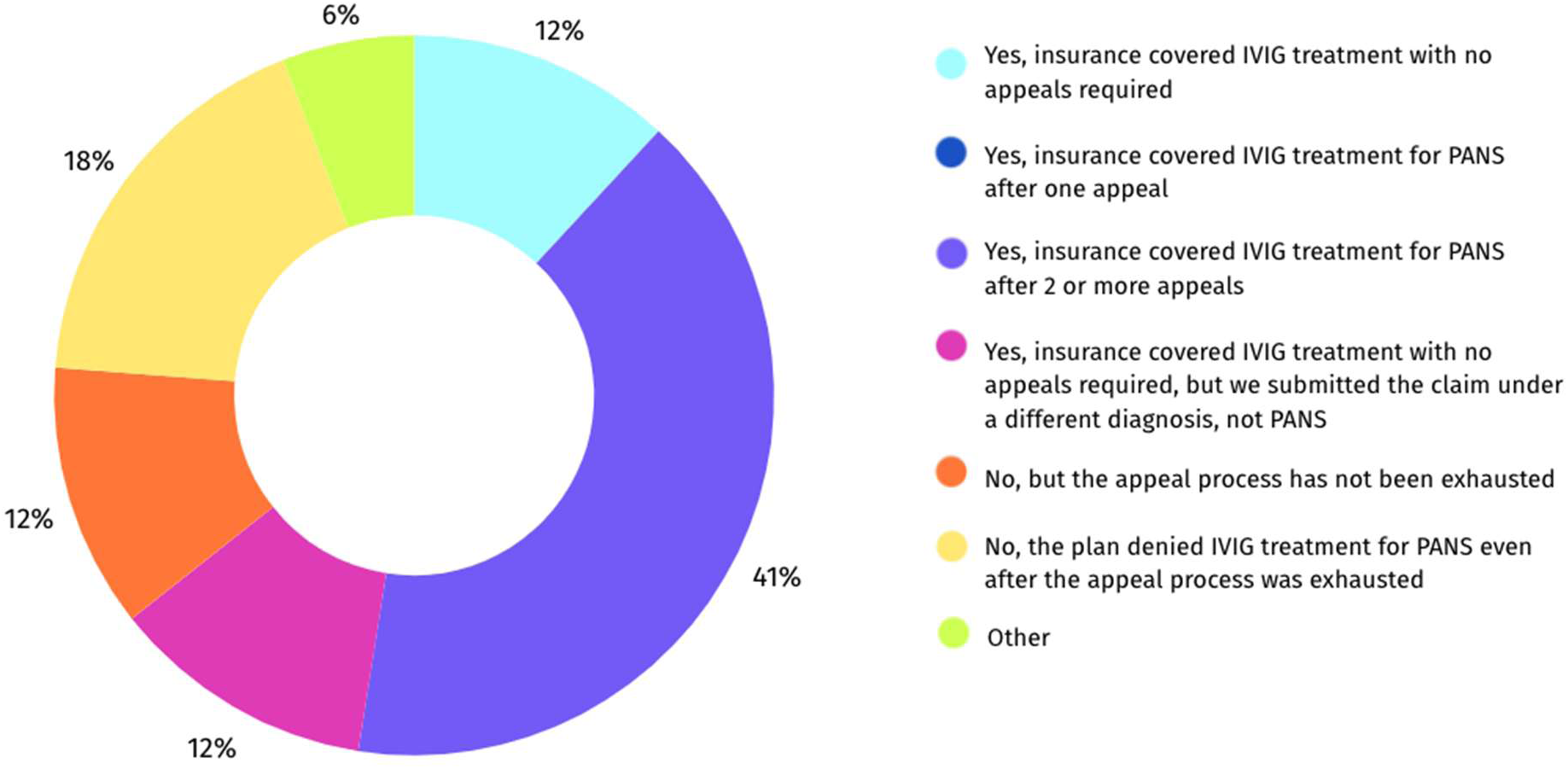
Insurance Coverage for IVIG Treatment (N=17)

As can be seen in Figure 2, those submitting IVIG claims for PANS alone were more likely than those submitting claims for comorbidities to wait at least a month before treatment could proceed; only 5% of those receiving IVIG for PANS were treated within a month of the order, and in 14% of cases, the waiting period was 9 months or more. In contrast, those submitting claims for conditions other than or in addition to PANS most commonly (38%) received treatment in less than a month. As described in the open-ended comments field to this question, one patient qualified for IVIG for a comorbid condition (selective antibody disorder) but was then disqualified because the doctor’s note mentioned PANS/PANDAS and did not re-qualify for coverage until all mention of PANS/PANDAS was removed. The burdensomeness of the process was articulated by several respondents, with one describing it as “brutal” and another stating that they “can’t count the number of hours spent.” Delays in treatment access—sometimes stretching months or years—were linked in the comments to worsening psychiatric symptoms, academic decline, and family stress.

**Figure 2.**
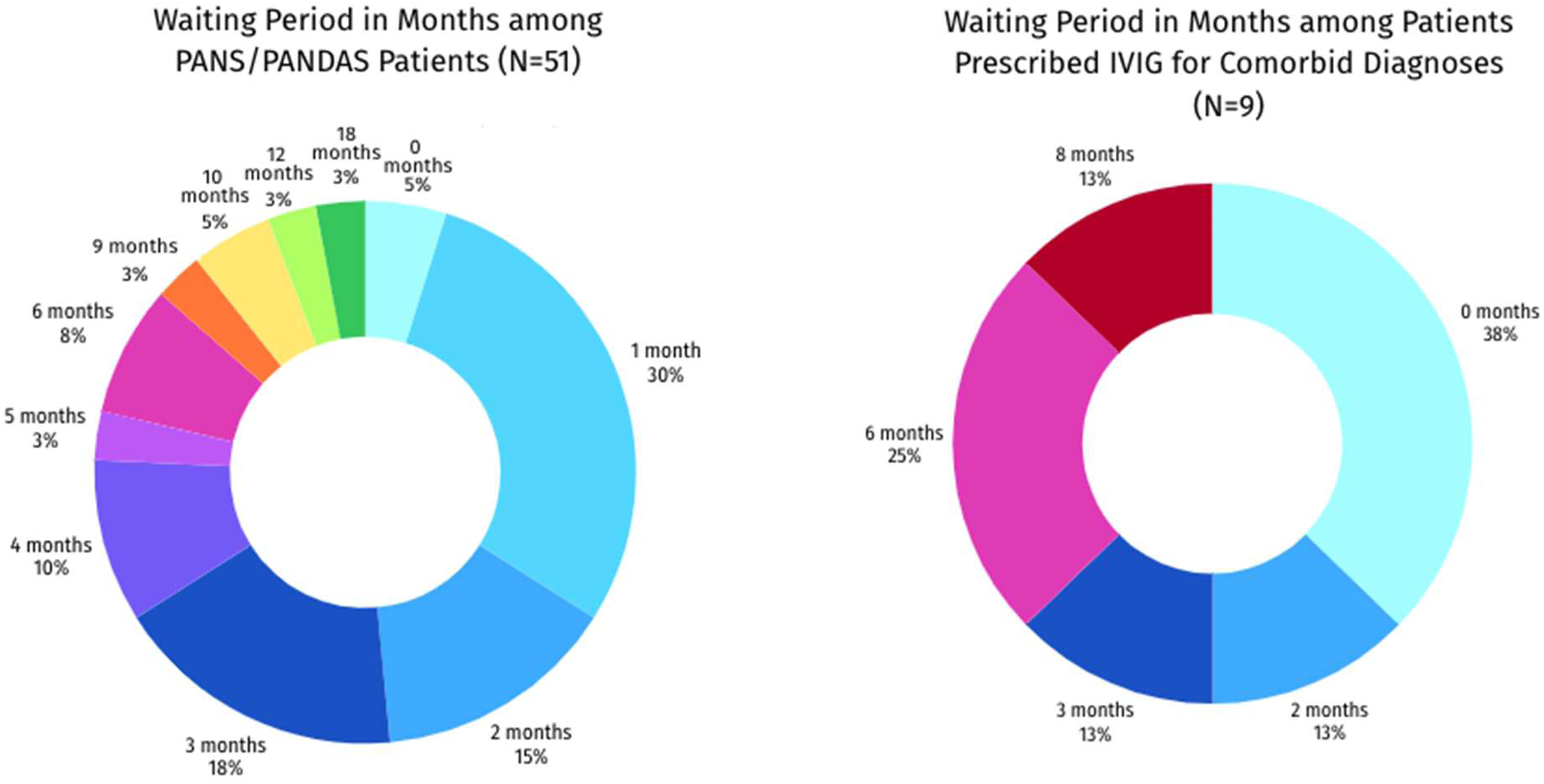
Waiting period between prescription and first IVIG treatment for PANS/PANDAS Alone vs. with Immune Deficiency and/or Autoimmune Diagnoses

### Financial Burden and Coping Strategies

As can be seen in Figure 3, paying for PANS treatment was associated with substantial financial strain for many families regardless of whether insurance assistance was obtained, and before IVIG was ordered in addition to after. According to the open-ended comments, some of this strain was associated with expenses not directly related to IVIG administration, primarily including the fees charged by specialists who are not part of insurance networks. Once IVIG was prescribed, however, average levels of financial strain diverged for families with versus without “substantial” insurance coverage (i.e. coverage of ≥70% of costs). For those without, financial strain often reached an extreme: during the six-month period beginning with the first IVIG administration, 33% of these, versus 13% of those with substantial insurance coverage reported strain levels at the very extreme of the scale (10 on a scale of 1 to 10). For those with substantial insurance coverage, the fraction expressing extreme levels of financial strain went down by nearly a factor of two once IVIG administration began, whereas for those without, this fraction nearly doubled.

**Figure 3.**
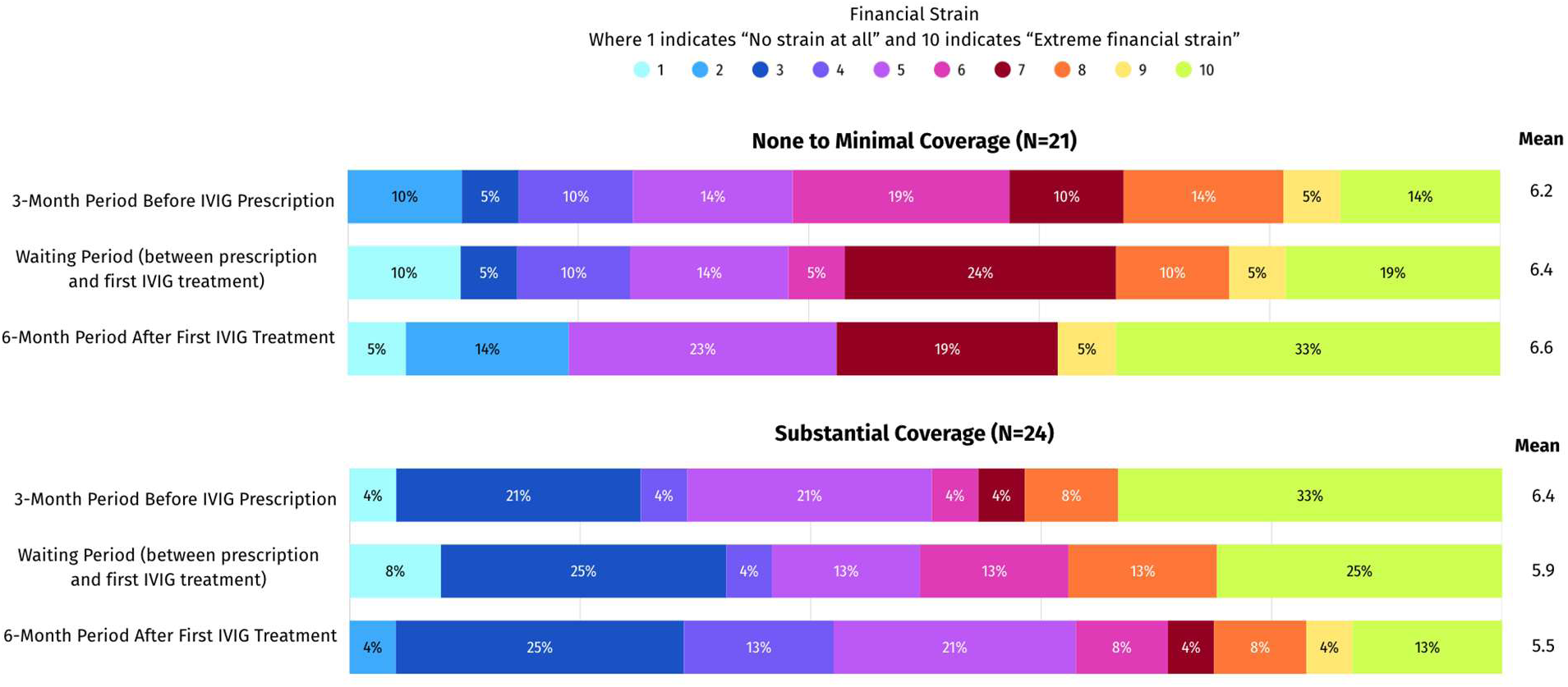
Level of Financial Strain Experienced Due to PANS Treatment, by Level of Insurance Coverage

**Table 3:**
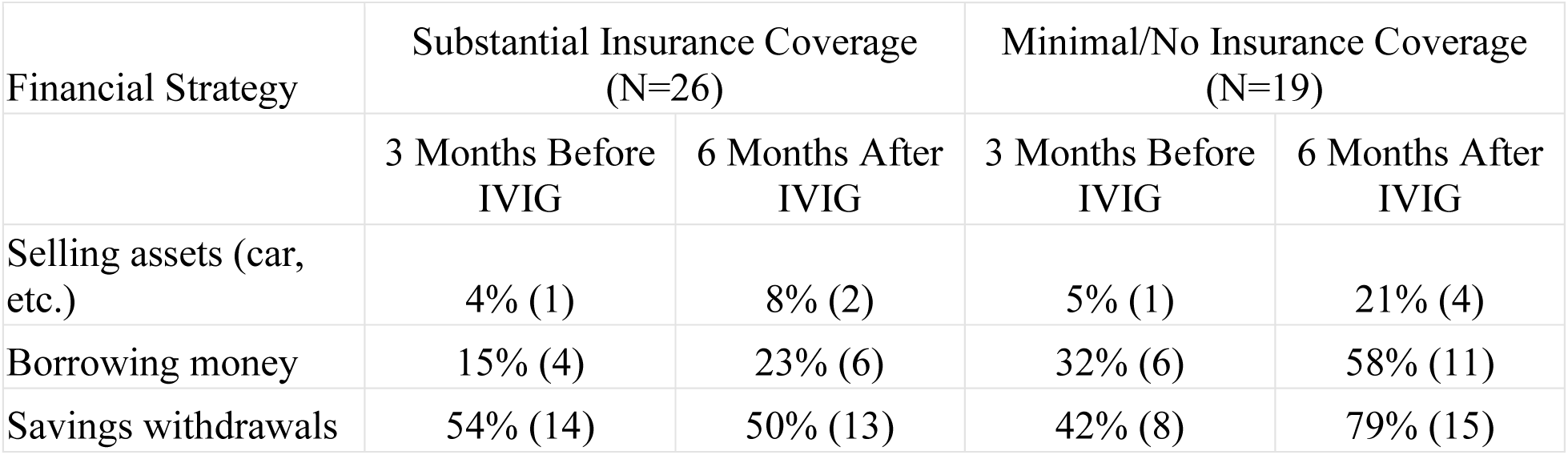
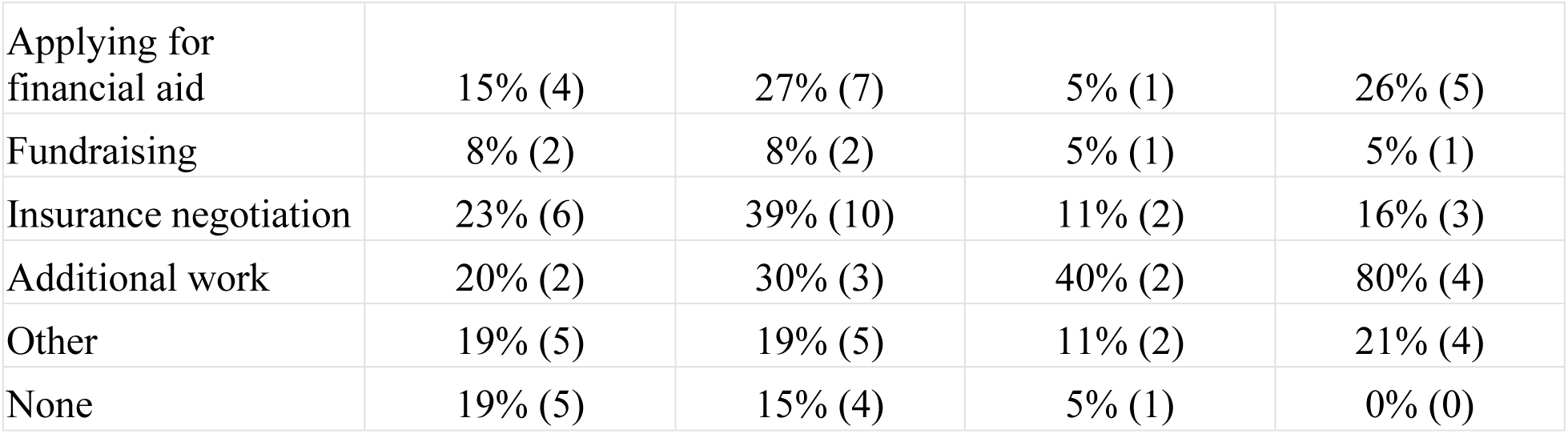
Financial Strategies Employed to Manage Costs of PANS Treatment by Insurance Status and Time Period.

With or without substantial insurance coverage, families commonly accessed savings to pay for PANS treatment; however, those with substantial insurance coverage were no more likely to do this during the period after PANS treatment started (50%) than they were before (53%), whereas among those without substantial coverage, the fraction withdrawing from savings increased from 42% before the IVIG order to 79% after. A similar pattern was seen for the fraction of families taking on additional paid work—which among those with substantial insurance coverage climbed from 20% before IVIG to 30% after, while among those without substantial coverage, climbed from 40% before to 80% after—as well as for the fractions who acquired loans and/or sold major assets to pay for PANS treatment. Among those without substantial insurance coverage, 58% acquired loans and 21% sold assets to pay for PANS care once IVIG had begun. Those families who ultimately obtained substantial insurance coverage were more than twice as likely as those who did not to have negotiated with insurers both before and after IVIG treatment was obtained (23-39% of those with substantial coverage, 11-16% of those without).

### Quality of Life

Across all measured domains, both patient and family/caretaker/cohabitant Quality of Life (QoL) demonstrated marked improvement following IVIG treatment, in comparison to the months preceding the IVIG order and the waiting period between order and first treatment. In terms of overall Quality of Life (Figure 4), means for both patients and family/caretaker/cohabitants (hereafter “family”) were very low prior to the IVIG order (2.3 and 2.6 respectively, on a scale of 1 to 10, with 1 indicating “exceedingly poor” and 10 indicating “exceedingly good”), with the majority endorsing ratings of 1 or 2. These remained essentially unchanged during the waiting period. After IVIG treatment, however, mean Overall Quality of Life ratings more than doubled, to 7.0 for patients and 5.7 for family/caretaker/cohabitants. Only 4% of the patients and 15% of family/caretaker/cohabitants remained at scores of 1 or 2, and nearly half of patients and a third of caregivers endorsed scores of 8 or above. Post-treatment family/caretaker/cohabitants ratings, but not patient ratings, were sensitive to the level of insurance coverage that had been obtained (Table 5).

**Figure 4.**
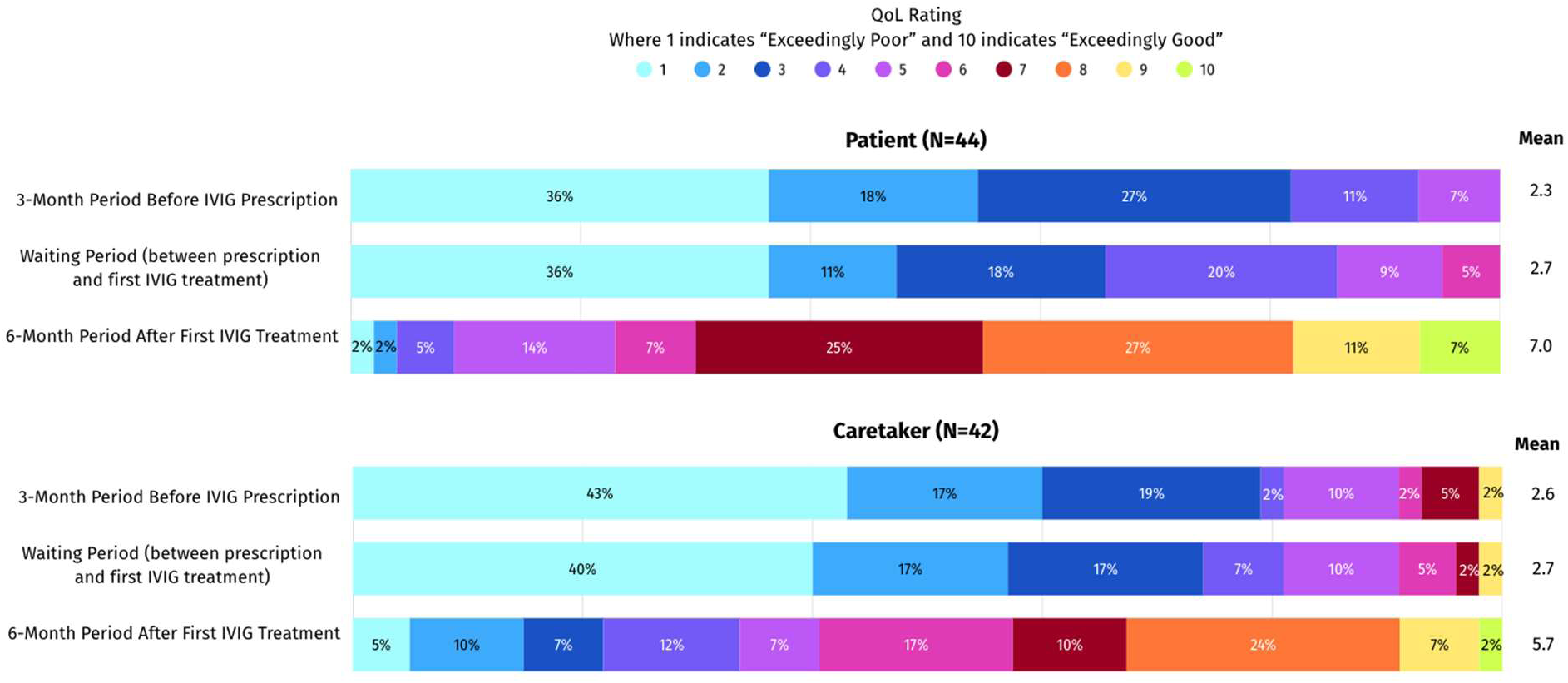
Quality of Life

**Table 5.** Patient and Family/Caretaker/Cohabitant Overall Quality of Life before and after IVIG Treatment, by Level of Insurance Coverage.

|  | Patient (N=44) |  |  | Family/Caretaker/Cohabitant (N=42-43) |  |  |
| --- | --- | --- | --- | --- | --- | --- |
|  | 3 months before IVIG order (mean) | Waiting period between order and dosing (mean) | 6-month period after first IVIG (mean) | 3 months before IVIG order (mean) | Waiting period between order and dosing (mean) | 6-month period after first IVIG (mean) |
| None to Minimal Insurance Coverage ($\leq 10\%$ of expenses covered) | 2.1 | 2.7 | 7.0 | 2.3 | 2.6 | 4.9 |
| Substantial Insurance Coverage ($\geq 70\%$ of expenses covered) | 2.8 | 2.7 | 6.4 | 2.5 | 2.7 | 7.0 |

With respect to specific domains, patients’ physical and emotional well-being prior to IVIG intervention was on average poor (mean=2.1, Table 6). Family/caretaker/cohabitant physical and emotional well-being was similarly low (mean=2.4) and both scores remained low during the waiting period (patient mean=2.5; family mean=2.5). However, during the six-month period post-IVIG, mean ratings more than doubled for both groups (patient mean=6.8; family mean=6.0). Scores for patients’ daily activities and interests were also low prior to the IVIG order (mean=2.8) and persisted low through the waiting period, but post-treatment mean scores again more than doubled to 6.8. Family ratings tracked closely, increasing from 2.5 and 2.6 pre-treatment to 6.1 after IVIG. Before IVIG treatment, impairment in academic and work performance was frequent among both patients (means 2.6 pre-order and 2.9 during waiting period) and family (means 4.0 pre-order, 3.9 waiting). During the six-month post-treatment follow-up, however, mean ratings yet again more than doubled to 6.8 for patients and 6.6 for caregivers. Social functioning and peer interactions followed a comparable trajectory.

**Table 6.** Patient and Family/Caretaker/Cohabitant Quality of Life before and after IVIG Treatment: Specific Domains.

|  | Patient (N=44) |  |  | Family/Caretaker/Cohabitant (N=42–43) |  |  |
| --- | --- | --- | --- | --- | --- | --- |
|  | 3 months before IVIG order (mean) | Waiting period between order and dosing (mean) | 6-month period after first IVIG (mean) | 3 months before IVIG order (mean) | Waiting period between order and dosing (mean) | 6-month period after first IVIG (mean) |
| Physical and emotional well-being | 2.1 | 2.5 | 6.8 | 2.4 | 2.5 | 6.0 |
| Social interactions | 2.6 | 2.8 | 6.2 | 3.0 | 3.2 | 5.7 |
| Educational/work performance | 2.6 | 2.9 | 6.8 | 4.0 | 3.9 | 6.6 |
| Daily activities and interests | 2.8 | 2.9 | 6.8 | 2.5 | 2.6 | 6.1 |

In the open-ended comment fields, respondents commonly reported severe lifestyle disruptions prior to treatment, with some caregivers forced to retire early (N=1), quit jobs (N=1), or risk unemployment due to missed work (N=1). Comments relating to the post-treatment period largely revolved around the notable improvements in patients’ school attendance and functioning.

## Discussion

Despite the very low prevalence of PANS and the limited fraction of even that small number prescribed IVIG, there are now laws in at least 15 U.S. states mandating insurance coverage for IVIG and other therapies in PANS, with similar legislation under consideration in 15 more. Yet until now, no systematic study has examined how families come to pursue IVIG, what they experience in attempting to obtain it, or what effects this treatment has on quality of life and functional outcomes for families. The current findings offer sobering insights into the real-world costs, delays, and emotional strain endured by families—and simultaneously suggest that meaningful benefit with this treatment is possible.

With respect to the initial pursuit of IVIG, a remarkable finding of this study was the extent to which families were influenced not only by medical professionals, but even more by their own personal research and the experiences of other families. Faced with a paucity of data regarding whether the impacts of IVIG could be expected to justify the often-profound financial sacrifices, parents appear to put considerable faith in the experiences of others in whose footsteps they follow, underscoring the critical role of peer-to-peer experience-sharing. Previous studies have documented the distressing journey many PANS families endure in obtaining a diagnosis and treatment^7,17,18^, as well as the extraordinary levels of caregiver stress suffered—above the “burnout” level^19^ and comparable to those suffered by caregivers of dementia patients^20^—but with the added burden that few people in the families’ lives can relate to their difficulties. The reliance of PANS caregivers on the lived experiences of peers who can relate closely to their own experiences may result from perceived weakness in the information and support available from other sources.

Also interesting was the wide range of specialists who ordered IVIG for the patients in our sample, reflecting the disorder’s lack of a specialty “home.” Although PANS presents primarily with psychiatric and neurological symptoms, its etiology and management are generally at least partly immunologic, making it difficult to situate within any one medical specialty. Not only does this fragmentation contribute to delays in diagnosis and effective treatment, but it may also lead to IVIG being ordered by practitioners who may not be expert in justifying its use to insurers or in identifying legitimate comorbid conditions for which obtaining coverage may be more straightforward. In our sample, IVIG had sometimes been prescribed for a comorbid immunologic or neurologic condition for which IVIG is generally covered, and as might be anticipated, the insurance approval process was more expeditious in these cases. Patients with PANS alone, in contrast, frequently reported long waiting periods, often extending beyond six months, before either receiving approval or abandoning hope. As illustrated by our quality of life findings, this prolonged the period of distinctively poor quality of life and function for both patient and family. Patients who experience prolonged delays before receiving effective treatment may also experience more negative outcomes over a period of years compared to those who are treated relatively promptly^15,16^.

Perhaps the most striking of our findings related to financial and emotional costs. Despite our survey respondents being relatively well-insured overall, many had resorted to exceptional measures to pay for IVIG treatment, and extreme levels of financial stress were reported by a large fraction. Remarkably, many respondents endorsed the extreme end of 10-point Likert scale, suggesting a level of distress so profound that a more stressful situation would be difficult to imagine. The substantial fraction of families who resorted to measures such as loans, asset sales, and additional employment is consistent with the gravity of this result.

Despite the challenges and sacrifices associated with obtaining IVIG treatment, the majority of respondents reported substantial improvements in function and quality of life post-treatment for both the child and the family. Interestingly, post-treatment quality of life metrics for most participants were not only higher than pre-treatment levels but often above the neutral range, with improvements including restored ability to attend school and/or work, improved well-being, greater enjoyment, and reintegration into family and social life. These results are generally consistent with those reported in published studies of IVIG in PANS. Of the two randomized controlled trials of a *single* round of IVIG in PANDAS, one showed significant benefit (45% reduction in OCD scale)^21^ and the other did not^22^. In the latter, non-responders were offered open label IVIg; a 49% decrease in OCD severity was seen in the 24 patients electing to receive this dose, raising the possibility that two rounds were needed to achieve impact in some of these patients. This possibility would be consistent with another study, which found that 85% of serious-severe or extreme PANS symptoms improved or resolved with IVIG but ∼3/4 of patients required 2 courses^23,24^. In more recent trials examining multiple doses of IVIg (3 to 6 courses every 3-4 weeks), significant improvements have been seen on OCD, tic, functional global impairment, and PANS symptomatology scales ^14,25,26^, and in one study, school absences after 3 courses dropped from 47% to 13% ^25^. In our study, most patients (98%) had received multiple courses of IVIG and improvements were substantial for both them and their parents; however, post-treatment quality of life ratings improved more for the patients than for their families and more for the substantially insured than for the underinsured, the discrepancy possibly reflecting the lingering financial and logistical burdens that families continued to bear even after the child’s health had improved.

This study had several limitations. Despite our efforts to recruit broadly and minimize selection bias, participants may have self-selected for strong opinions or extreme experiences. The retrospective nature of the data also raises the possibility of recall bias. Moreover, insurance coverage laws are evolving rapidly, and the experiences reported here may not reflect current conditions in all U.S. states, let alone outside of the U.S. Ongoing investigation is needed to determine whether recent legislative mandates in the U.S. are resulting in meaningful improvements in access, as well as improved patient and family clinical and quality of life outcomes. Research into treatment-access journeys of families outside of the U.S., who face quite different situations with respect to payment for healthcare, would also be of interest.

Additional clinical trials of this rare disorder are also needed, not only to better quantify the benefit-cost ratio of IVIG but also to better ascertain whether and in what patients more cost-effective treatments, for example traditional DMARDS (disease-modifying anti-rheumatic drugs), antibiotics, and/or anti-inflammatory medications, may be sufficient to obtain the same level of clinical benefit.

## Data Availability

All data produced in the present study are available upon reasonable request to the authors.

